# Emergence of the E484K Mutation in SARS-CoV-2 Lineage B.1.1.220 in Upstate New York

**DOI:** 10.1101/2021.03.11.21253231

**Authors:** Emil Lesho, Brendan Corey, Francois Lebreton, Ana C. Ong, Brett E. Swierczewski, Jason W. Bennett, Edward E. Walsh, Patrick Mc Gann

## Abstract

Ongoing surveillance detected a SARS-CoV-2 B.1.1.220 variant carrying the E484K substitution in four patients from a hospital network in upstate New York. Patients reported no travel history and shared no obvious epidemiological linkage. A search of online databases identified 12 additional B.1.1.220 with E484K, all of which were detected in New York since December 2020. Detailed genomic analyses suggests that the mutation has emerged independently in at least two different B.1.1.220 strains in this region.

## Text

Severe acute respiratory syndrome coronavirus 2 (SARS-CoV-2), the etiological agent of coronavirus disease 2019 (COVID-19), has caused a global pandemic that is reshaping society. As part of an unprecedented global response from the scientific community the development and rapid deployment of multiple antibody-based countermeasures that target the viral spike protein has proceeded at an incredible pace.

Over the past six months, the emergence of three variants with mutations in the spike protein has raised serious concerns about the durability of the current suite of vaccines and immunotherapies. The variants, which are colloquially referred to as the United Kingdom (B.1.1.7 using the Pangolin nomenclature^1^), South African (B.1.351), and Brazilian (P.1) variants^2-4^, have been increasingly identified worldwide^5^. The first variant (B.1.1.7) was identified in September 2020 and carries a N501Y mutation in the receptor-binding domain (RBD) of the spike protein^4^. Critically, recent studies have suggested this mutation potentially increases transmission and virulence^6,7^. Shortly thereafter, this same mutation was identified in B.1.351 and P.1^2,3^. Both variants also carried an additional substitution (E484K) in the spike that can increase resistance to neutralization by many monoclonal antibodies (mAb), while most convalescent sera and mRNA vaccine-induced immune sera show reduced inhibitory activity^8-11^. Notably, interim results from the Novavax adjuvented spike protein nanoparticle COVID-19 vaccine trial indicated an efficacy of 85.6% against B.1.1.7 infection (versus 95.6% for the original strain), but just 49.4% to 60% in a South Africa Phase 2b trial in which the B.1.351 strain was detected in the majority of COVID-19 events^12^.

In the United States, there have been 3,037 cases with the B.1.1.7 variant identified in 49 jurisdictions, 81 cases with B.1.351 (20 jurisdictions), and 15 cases with P.1 (9 jurisdictions) as of this writing (March 2021)^13^. In addition, several regional variants have been identified, such as the 20C-US lineage in the Midwest^14^ and the B.1.2 lineage carrying the Q677P substitution in New Mexico and Louisiana^15^. However, these lack the E484K and N501Y mutations. In contrast, the recently described B.1.536 variant in New York City carries E484K^16^. Notably, this variant has been spreading at an alarming rate since November and by February 15^th^ 2021 accounted for 12.3% of all cases in this catchment area. Similar to previous studies with variants carrying E484K, antibody neutralization by mAbs REGN10933, CB6, and LY-CoV555 are either impaired or abolished with this variant^16^. Similarly, the neutralizing activities of convalescent plasma or post-vaccination sera were lower by 7.7-fold and 3.4-fold, respectively^16^. These data illustrate the alarming rise of this mutation, which has now been detected in 88 of 886 lineages (as of this writing), with various degrees of frequency^5,17^.

In June 2020, a pilot study to investigate SARS-CoV-2 was initiated between the Multi-drug resistant organism Repository and Surveillance Network (MRSN) and the Rochester Regional Health System; a network consisting of 8 acute care hospitals, 9 urgent care centers, and 6 long-term care facilities in St Lawrence County and the 9-county Finger Lakes Region of NY. Every hospital-based laboratory in the network is capable of testing for COVID-19 infection and to date 438,107 samples have been tested with 25,468 distinct patients testing positive. For this study, samples were provided by five hospitals in the Finger Lakes region. RNA is extracted from SARS-CoV-2-positive nasopharyngeal swabs and transferred to the MRSN for sequencing (See Methods). To date, >150 samples spanning April 2020 to February 2021 have been analyzed, with 14 different Pangolin lineages identified (manuscript in preparation). Though notable mutations were found in the spike proteins from these earlier samples, including D614G, S673T, Q677H, N679K, and P681H, the E484K and N501Y substitutions were not identified.

The most recent shipment consisted of 20 samples from 19 patients collected between January 27^th^ and February 7^th^, 2021. Upon analysis, the E484K substitution was detected in five samples from one male and three female patients. The average age was 85 years (range 74-92) and, despite advanced age and multiple co-morbidities (mean CALL score = 10)^18^, all recovered. Three patients had mild courses with no specific anti-SARS-CoV-2 treatments. One patient received Remdesivir and Dexamethasone and did not require intensive care or endotracheal intubation. Notably, three patients lived in separate assisted living facilities with no known connections and the fourth resided at home. All patients reported no significant travel in the preceding 6 months.

The five strains were assigned to Pangolin Lineage B.1.1.220 (NextStrain clade 20B; **Figure 1A**), a globally distributed lineage that has also been identified in 2-7% of COVID-19 cases in the North-East US (**Figure 1B**). In addition to the E484K substitution, the strains carried the characteristic 4 amino acid substitutions that define this lineage when compared to the Wuhan-Hu-1 SARS-CoV-2 reference genome (NCBI GenBank Acc. MN908947), namely R203K, G204R, P314L and D614G. This latter substitution has become the most prevalent mutation world-wide and studies suggest it increases viral transmissibility and results in higher viral loads ^19,20^. To better understand the relationship between the 5 samples we performed a whole genome high-resolution single nucleotide polymorphism (SNP) based comparison. The two samples from the same patient were genetically identical (0 SNPs), but the remaining samples were separated by 1-4 SNPs.

**Figure 1.**
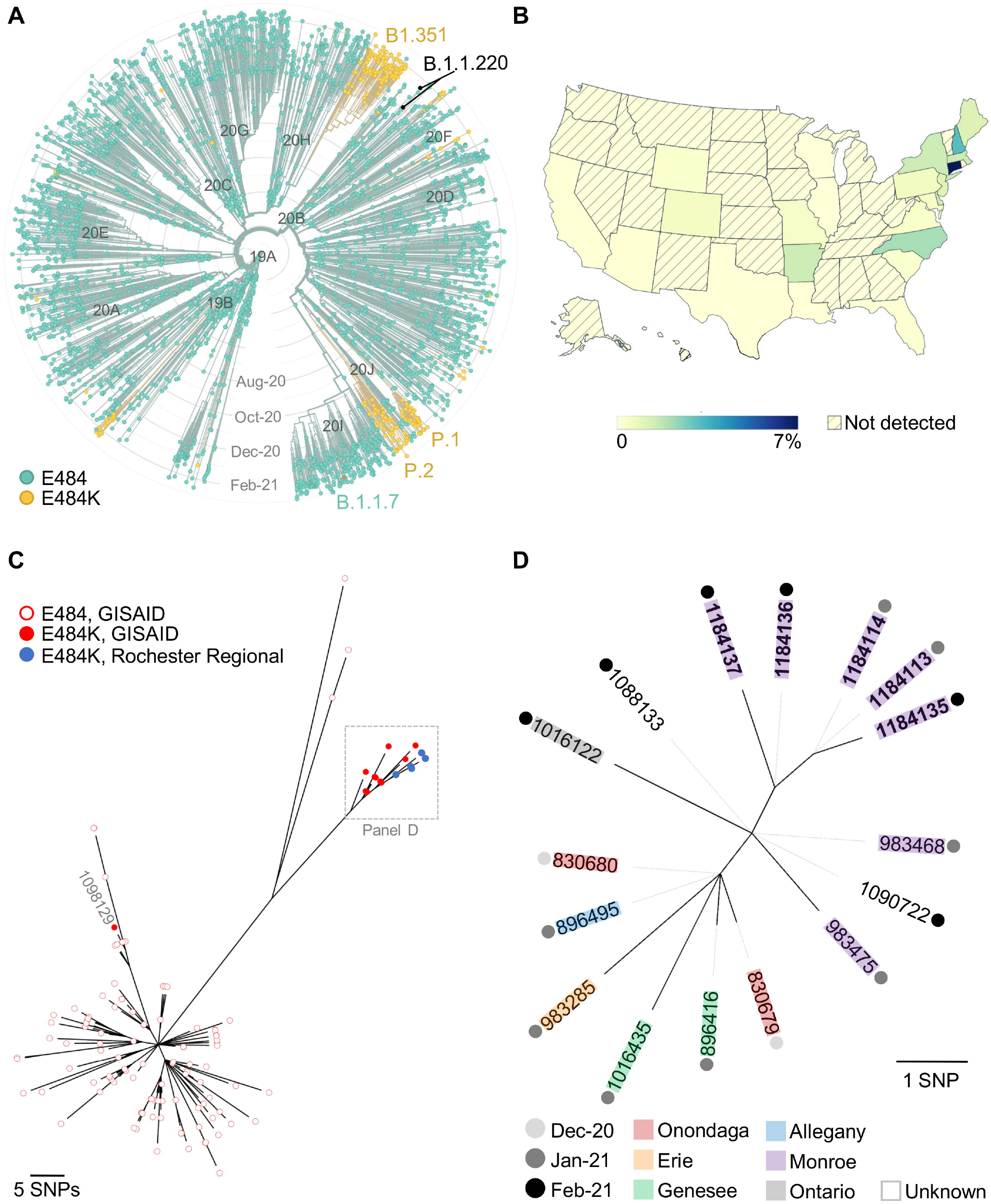
Emergence of the E484K mutation in B.1.1.220 SARS-CoV-2 isolates from Upstate New York. **(A)** Time-scaled, global phylogeny of 3,960 SARS-CoV-2 genomes sampled between Dec 2019 and Mar 2021 (modified from Nextstrain^27^ dashboard). A full table of acknowledgements for the data included in this analysis is available in the supplement (Table S2). Major clades (Nextstrain nomenclature), variants (literature) and lineages (Pangolin) are indicated. Strains are colored based on the presence of a glutamic acid (E, teal) or lysine (K, orange) at position 484 of the spike protein. **(B)** Color-gradient map of the USA showing the estimated percentage of sequenced SARS-CoV-2 genomes that were assigned to Lineage B.1.1.220 since first detection (modified from Outbreak.info^17^ dashboard). **(C)** SNP-based, unrooted phylogram of B.1.1.220 variants collected from New York State between October 2020 and March 2021 (n = 129). Empty circles indicate strains with Glutamic acid (E) at residue 484 of the spike protein while filled circles indicate those with Lysine (K) at that position. Genomes obtained from GISAID^26^ (Table S1) are shown in red. Genomes newly described in this report are shown in blue (note that only four out of five circles are visible as two samples from a single patient were identical). **(D)** SNP-based, unrooted maximum-likelihood phylogeny of sixteen monophyletic B.1.1.220 strains carrying the E484K mutation. Both comparator (Table S1) and newly sequenced isolates (bold font) are labeled using their GISAID Accession ID. Where known, the county and collection date are indicated. For both panels C and D, the number of SNPs separating each strain can be inferred from the respective scale.

Analysis of the five variants in the open data software at MicroReact^21^ identified 6 additional B.1.1.220 variants with the E484K substitution. All were deposited by the Wadsworth Center (Albany, NY), a sentinel institute of the New York Department of Health. Further comparative analysis with 124 B.1.1.220 sequences from the New York region deposited at GISAID identified an additional 6 variants with this mutation. Surprisingly, a phylogenetic analysis suggested that the E484K substitution has arisen independently at least twice within Lineage B.1.1.220 in the New York region, as evidenced by the significant genetic separation (25 to 29 SNPs) between a monophyletic cluster of 16 samples from Central and Western New York and sample 1098129 from the New York Metropolitan area (**Figure 1C, Table S1**). Zoomed-in analysis of the 16 monophyletic strains revealed high-genetic relatedness (0-7 SNPs) and further suggested that this B.1.1.220 E484K variant has just recently emerged and is rapidly spreading across New York State with isolates being collected from at least 6 distinct counties between December and February 2020 (**Figure 1D**). Importantly, as of this writing (March 2021) the only B.1.1.220 variants shown to harbor the E484K mutation have been isolated in New York.

To the best of our knowledge, this is the first report of a Lineage B.1.1.220 bearing the important E484K mutation. The observed mutational convergence within the relatively small subset of Lineage B.1.1.220 genomes from New York State further indicates that codon S/484 is evolving under a strong degree of positive selection. Though not supported statistically, it is notable that no B.1.1.220 variants were detected in our surveillance effort prior to December 2020 when it suddenly comprised 25 % of the late January and early February samples. However, as of this writing (March 2021), only 146 B.1.1.220 genomes from NY have been uploaded to GISAID, with the majority (n = 87) collected since January 1^st^, 2021; additional B.1.1.220 sequences from this region are required to better understand the current distribution and apparent emergence of this concerning variant. It will also be important to assess how the E484K mutation affects protection by vaccines against this specific variant that lacks the other mutations in the B1.351 and P1 strains. As the vaccine roll-out gathers pace across the United States, it is imperative that every means necessary are employed to rapidly identify the emergence of variants-of-concern and immediate action taken to limit their spread.

## Methods

### Sequencing of SARS-CoV-2 samples

Viral RNA was extracted from nasopharyngeal swabs using the QIAamp Viral RNA Mini Kit (Qiagen, 52906). RNA was converted to cDNA and enriched for SARS-CoV-2 using the QIAseq SARS-CoV-2 Primer Panel (Qiagen, 333896). Libraries were constructed using the KAPA HyperPlus kit (Roche, KK8514), quantified with the KAPA Library Quantification kit (Roche, KK44), pooled, denatured and diluted to 14 pM. The diluted sample was sequenced on an Illumina MiSeq sequencer using a MiSeq v3 (600 cycle) Reagent Kit (Illumina, MS-102-3003).

### Bioinformatic Analysis of SARS-CoV-2 genomes

Miseq read data was trimmed for adapters and low quality sequence using BBduk v38.79 (https://sourceforge.net/projects/bbmap/). Trimmed reads were then mapped to the Wuhan-Hu-1 SARS-CoV-2 reference genome (NCBI GenBank Acc. MN908947) using Snippy v4.4.5 (https://github.com/tseemann/snippy). BAM files were imported into Geneious^22^ where variants identified by Snippy were manually curated/confirmed. Nucleotide alignments were performed with Muscle^23^ followed by trimming of ambiguous/under-represented sequence at the 5’/3’ ends of the alignment in Geneious. Genome-based, maximum likelihood phylogenies were produced with RAxML-ng (model GTR+Gamma, 50 parsimony, 50 random) ^24^. Best scoring phylogenetic trees were selected and edited using iTOL^25^.

### Use of GISAID, Microreact, NextStrain, Outbreak.info and Pangolin Resources

Lineage assignment was performed using the Pangolin online web application^1^. Identification of additional B.1.1.220 lineage samples with and without the E484K mutation was performed initially with Microreact^21^ and later with GISAID^26^. An acknowledgement of the originating and submitting laboratories providing the SARS-CoV-2 comparator genomes used in this study can be found in the Supplemental Material (**Table S1** and **S2**).

The time-scaled analysis of SARS-CoV-2 genomes (Fig. 1A) was modified from the Nextstrain^27^ SARS-CoV-2 global phylogeny (https://nextstrain.org/ncov/global) accessed on March 6^th^, 2021. This build contained 3,960 SARS-CoV-2 genomes sampled between Dec 2019 and Mar 2021 and collected from: Africa (n = 588), Asia (n = 671), Europe (n = 783), North America (n = 845), Oceania (n = 489) and South America (n = 594). Full details on bioinformatic processing are available on the Nexstrain Github page (https://github.com/nextstrain/ncov).

The USA map of the estimated Lineage B.1.1.220 prevalence (Fig. 1B) was created and edited from the Outbreak.info^17^ dashboard accessed on March 7^th^, 2021.

## Data submission

The sequences of the five strains described in this paper have been uploaded to GISAID with Accession ID (virus name): EPI_ISL_1184113(hCoV-19/USA/NY-RRHS-101/2021), EPI_ISL_1184114 (hCoV-19/USA/NY-RRHS-102/2021), EPI_ISL_1184135 (hCoV-19/USA/NY-RRHS-114/2021), EPI_ISL_1184136 (hCoV-19/USA/NY-RRHS-115/2021), and EPI_ISL_1184137 (hCoV-19/USA/NY-RRHS-120/2021).

## Acknowledgements

We are deeply indebted to the clinical and research laboratories of the Rochester Regional Health System and ACM Global Laboratories, especially Maria Formica, Julie Freed, Chalece Holland, Jean Campbell, and Julia Nary.

This study was partially funded by the Defense Health Program (DHP) Operations & Maintenance (O&M). Material has been reviewed by the Walter Reed Army Institute of Research. There is no objection to its presentation. The opinions or assertions contained herein are the private views of the authors and are not to be construed as official, or reflecting the views of the Department of the Army or the Department of Defense. Some authors (Bennett, Mc Gann, Swierczewski) are employees of the U.S. Government and this work was prepared as part of their official duties.

